# The efficacy of an online behavioural intervention for improving dietary habits with a focus on self-compassion, goal-setting and self-monitoring: A randomised controlled trial

**DOI:** 10.1101/2023.04.18.23288716

**Authors:** Hania Rahimi-Ardabili, Rebecca Charlotte Reynolds, Nicholas Zwar, Nancy Briggs, Lenny R. Vartanian

**Author notes:** **Corresponding author:** Dr Hania Rahimi-Ardabili will handle correspondence at all stages of refereeing and publication, also post-publication. Hania Rahimi-Ardabili was at the School of Population Health when the research was conducted.

## Abstract

**Issue addressed:** To examine the efficacy of an online intervention that combined self-compassion with goal-setting and self-monitoring to improve dietary habits.

**Methods:** Australian adults with overweight and obesity were randomly assigned to the intervention or control group. A 12-week online nutrition intervention that included self-compassion, goal-setting and self-monitoring compared to a control group who received nutrition information only. Measures assessed before and after the intervention included primary outcomes of self-compassion; eating pathology; depression, anxiety and stress; and dietary intake, and a secondary outcome of anthropometry (weight and body mass index). Analyses were completed by a series of 2 (time: pre- and post-intervention) by 2 (group: intervention and control) mixed analyses of variance (ANOVAs) using the ‘intention-to-treat’ approach.

**Results:** 117 people (73 intervention, 44 control) participated; of those, 74 participants (35 intervention, 39 control) completed the intervention. The analysis of all 117 participants showed that some aspects of dietary intake improved in the intervention group but not in the control group (i.e. time*group interaction), including a decrease in energy intake (Coefficient=2139.23, p<0.001 vs Coefficient=169.29, p=0.82), carbohydrate intake (Coefficient=56.22, p=0.006 vs Coefficient=-9.43, p=0.71); and a marginal reduction (*p*s=0.06) in saturated fat intake and improvement in fruit scores.

**Conclusions:** The findings suggest that the intervention could improve dietary habits. Further studies are needed to confirm these findings, examine the efficacy of the intervention over a longer period, and determine the mechanism underlying these changes.

So what? Online interventions that contain self-compassion, goal-setting and self-monitoring have the potential to promote healthy dietary habits.

## INTRODUCTION

A growing body of literature emphasises the importance of considering psychological factors related to overweight and obesity in weight management interventions^1^. Psychological and behavioural factors, such as body dissatisfaction and disordered eating, might contribute to poor outcomes^2^. Self-compassion (i.e. a tendency to treat oneself with kindness in times of suffering^3^) is an approach that might be useful in this context.

Previous studies have reported that self-compassion is beneficial for body-acceptance and self-acceptance^4,5^ and for decreasing disordered eating^6,7^. Self-compassion can also be directly beneficial for improving dietary habits. Evidence suggests that people high in self-compassion might have greater behaviour change skills (e.g. goal-setting, self-monitoring), meaning that they might have more intrinsic motivation to pursue their goals, and show less negative reaction towards minor goal setbacks compared to people low in self-compassion^8,9^. Negative reactions to minor failures and lack of motivation^10^ are considered factors that contribute to health goal abandonment. Studies on the self-compassion intervention for dietary behaviour is promising but preliminary, often with methodological limitations such as lack of control arms^14-16^ or conducted on different participant groups (e.g. clinically diagnosed patients) ^11,12 13^ that make it difficult to generalise to people with overweight or obesity.

Another factor that could be relevant to the effectiveness of programs aimed at improving dietary habits is the availability of resources which can be common barriers for participating in programs for dietary habit change^17^. A potential solution to this problem is the use of online interventions^18^, which could also provide facilities such as a platform for self-monitoring, delivering reminders, and providing in-time feedback on progress^19^ to increase participants’ engagement with self-monitoring compared to paper-based self-monitoring^20^.

The current study sought to combine self-compassion and online intervention approaches to improve dietary habits. In particular, the current randomised controlled trial (RCT) investigated the efficacy of an online 12-week intervention that combined self-compassion with goal-setting and self-monitoring to improve dietary behaviour in people with overweight or obesity compared to the control group. This study also aimed to explore whether the effects of the intervention on the outcomes were explained by changes in self-compassion.

## METHODS

### Study design

The study was a 12-week parallel RCT. Ethics approval for this study was obtained from an ethics committee [information removed for blinding]. The trial was registered with a clinical trial registry [information removed for blinding].

### Participants

The target population was individuals with overweight or obesity living in Australia, and participants were recruited from July 2017 until January 2018. Participant inclusion criteria were: aged 18 - 55 years; BMI of 25 - 40 kg/m^2^; able to run internet browser for at least one hour per week; able to read and write English. The exclusion criteria were: taking any weight-loss medications or previous use of weight-loss medications during the past six months; currently using medication which is associated with substantial weight gain; suffering from or having a history (in the last five years) of any major medical illness; pregnancy or lactation; current participation in any other nutrition or weight loss program; currently smoking; and weight loss of more than 4.5 kg (10 pounds) during the past six months.

### Sample size

Power analysis was based on the effect size detected in an initial pilot study that was conducted to assess the feasibility and efficacy of the current intervention^14^ for self-compassion outcome (measured by the Self-Compassion Scale [SCS]). The sample size was calculated to detect the effect size of Cohen’s *d*=0.61 for the changes between two groups with a power of 80% and a confidence interval (CI) of 95%. A final sample size of 45 was required in each group, which was increased by 30% to take attrition into account. Therefore, a sample size of 60 in each group was aimed for (i.e. a total sample of 120).

### Recruitment and randomisation

Participants were recruited from the general population across all Australia online using various approaches of email, online newsletters and social media. Participants who finished the study were entered to a prize draw to gift vouchers. A random sequence of 120 numbers was generated in two balanced - columns using the online website Random.org. The method of sealed envelopes suggested by Schulz and Grimes^21^ was used for allocation concealment, which seeks to prevent selection bias. The study was a single-blind trial: the lead investigator ([information removed for blinding]) was aware of what group participants were allocated to (after the screening was completed). After randomisation, online informed consent was sought from participants. Participants were not told which group they were allocated to but were informed that there were two different conditions.

### Data collection

The whole process of data collection was performed online. Participants completed pre- and post-intervention assessments of their level of self-compassion; eating pathology; the level of depression, anxiety and stress; dietary intake; and anthropometry. Demographic data, such as age, were collected at baseline only.

### Self-Compassion Scale

The Self-Compassion Scale (SCS) is a 26-item self-reported measure designed to assess typical thoughts, emotions and behaviours associated with different components of self-compassion^22^. The self-compassion scale consists of six subscales: Self-Kindness, Self-Judgment, Common Humanity, Isolation, Mindfulness and Over-Identification. Responses are made on a five-point scale from 1 (Almost never) to 5 (Almost always). Subscale scores are computed as the mean of items in subscales ^22^. Internal consistency reliability for the SCS was good, with Cronbach’s alpha between 0.77 and 0.88 for subscales.

### Eating Disorder Examination Questionnaire

The Eating Disorder Examination Questionnaire (EDE-Q) is a 28-item questionnaire that asks about maladaptive eating behaviours over the previous four weeks^23^, and provides two types of data: First, it generates a frequency of occurrence of the main behavioural traits of eating disorders such as binge eating (six questions). Second, it has subscale scores that provide the severity of eating-related psychopathology^23^. These items are responded to on a scale that ranges from 0 (No days) to 6 (All days). The four subscales are: Restraint, Eating Concern, Shape Concern and Weight Concern. The score for each subscale is obtained by calculating the mean of all items for that subscale. A Global score for overall eating pathology, is obtained by averaging the four subscale scores. Higher EDE-Q scores reflect a greater severity of eating psychopathology. In the current study, internal consistency for EDE-Q Global was good (Cronbach’s alpha=0.89), and for its subscales, alpha values ranged from 0.72 to 0.89.

### Depression Anxiety and Stress Scale-21

The Depression Anxiety and Stress Scale-21 (DASS-21) is a 21-item self-administered instrument assessing psychological distress^24^. It is composed of three subscales: Depression, Anxiety and Stress. Respondents indicate the extent to which they experienced negative emotional states over the past week, ranging from 0 (Did not apply to me) to 3 (Applied to me very much) ^24^. To attain a score for each subscale, the ratings for the subscale items are summed. Internal consistency was also good for all DASS subscales (Cronbach’s alpha of 0.87, 0.80 and 0.88, respectively).

### Dietary assessment

Dietary intake was evaluated using two different methods. A validated and web-based self-administered 24-hour dietary recall (INTAKE24)^25^ for nutrient-based assessment, and the Healthy Eating Quiz (HEQ) for food-based assessment^26^. INTAKE24 asks about food consumption in the last 24 hours or the previous day^27^. HEQ reflects the frequency of foods (and drinks) or food groups consumed in a particular period and allocates points based on consumed food types and/or frequency^28^. The HEQ uses 70 items from the validated Australian Eating Survey Food Frequency Questionnaire (AES FFQ)^26^. The highest score for HEQ is 73, and higher scores reflect better alignment with the Australian Dietary Guidelines^26^.

### Anthropometry

Weight and height were obtained via self-report. Participants were asked to measure their weight and height with their personal weighing scales and measuring tapes based on the instructions^29^ provided to them before completing the surveys.

### Intervention

The intervention materials and the online application used in this study was developed based on findings from an initial pilot study that assessed the acceptability and efficacy of the self-compassion, goal setting and self-monitoring intervention in people with overweight and obesity^14^. The formatting of the information for this study was refined based on the feedback received from pilot study participants to be more concise and engaging. Some of the self-compassion information was modified to cover topics that are directly related to body image and eating behaviours. More information about the pilot study is available elsewhere^14^.

### Development of the online application

An online application was developed to provide a user-friendly and accessible tool for participants to set and monitor goals. The basic behaviour-change principles such as goal-setting, self-monitoring, problem-solving and barrier identification to facilitate dietary changes^30^ were used for the development of the application. The application also provided feedback on participants’ performance, sent reminders when the participants did not log into their accounts for 24 hours and was compatible with any device (e.g. smartphone, laptop).

The investigators designed the application to promote goal-setting, according to Locke and Latham’s^31^ theory, to include attributes that facilitate goal attainments, such as specificity and proximity. For this purpose, the application required participants to select goal types, enter their goal, and set how often to accomplish a goal per week). NoMoss (Co Pty Ltd, Sydney, Australia) designed and created the application based on ongoing feedback provided by the lead investigator (). Example screenshots from the application are provided in Figure 1. The application was pilot tested using the 10-item System Usability Scale^32^. Thirteen volunteers rated items such as ‘*I thought the system was easy to use*’ on a five-point Likert scale ranging from zero (‘Strongly disagree’) to four (‘Strongly agree’).. The median score for the application was 75, within the acceptable range of ≥70^33^. After pilot testing, any errors were fixed, and the application was also modified to improve the application’s usability (e.g. changes in wordings, display colour and fonts).

**Figure 1.**
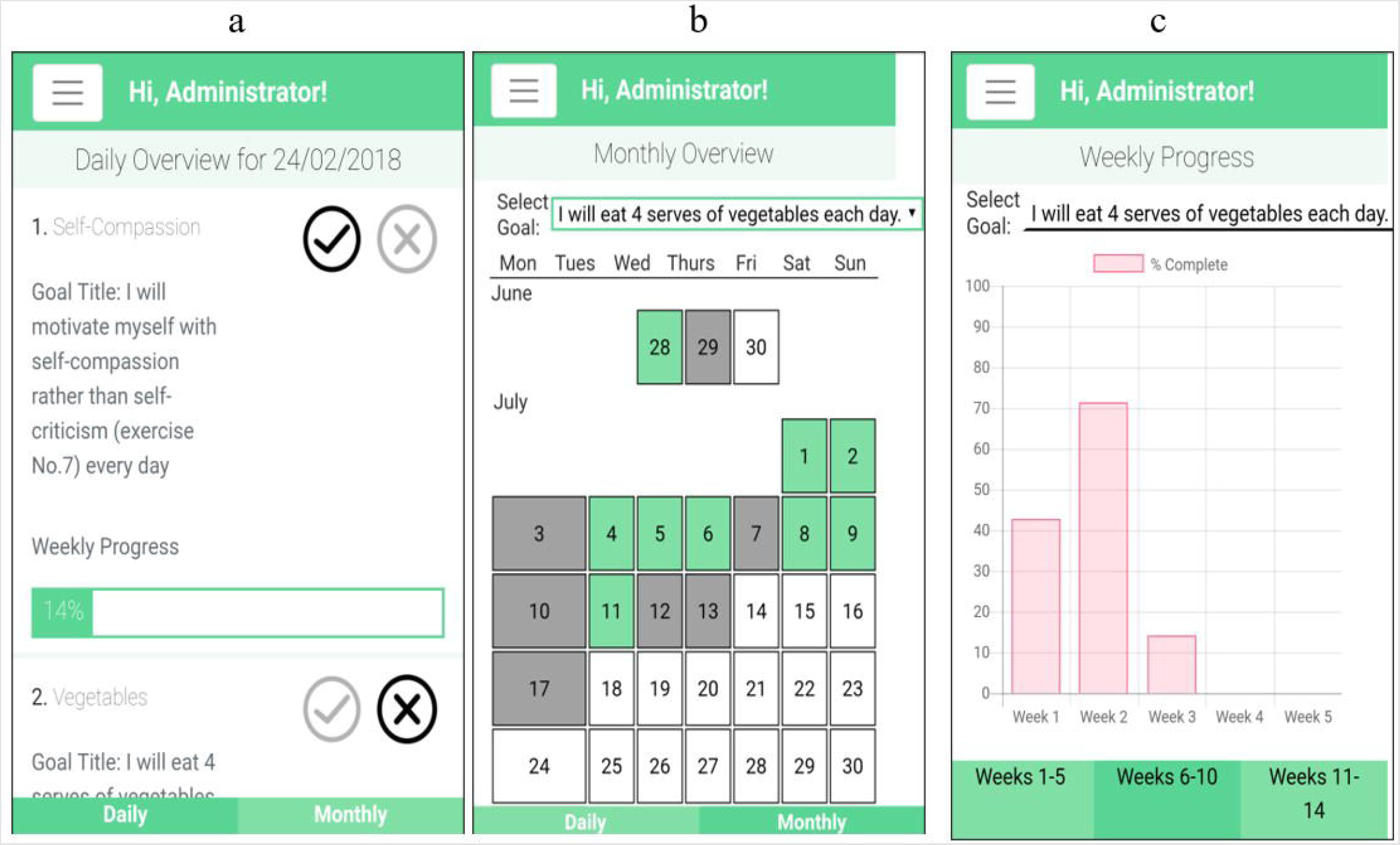
Online application’s ‘Daily Overview’ (a), ‘Monthly Overview’ (b) and ‘Weekly Progress’ (c) screens

### Overview of the study protocol

Participants then received an email with a link to the study website every four weeks (three emails in total: Week Zero, Week Four and Week Eight). The website provided information about nutrition and self-compassion. The initial round of information (Week Zero) also contained orientation and instructions.

Participants in the control group received an email every four weeks that provided only a brief form of standard nutrition information in PDF format (three emails in total: Week Zero, Week Four and Week Eight). The nutrition information provided to the control group was similar to the core part of the nutrition information provided to the intervention group. This group did not receive any information on goal-setting or self-compassion, nor did they have access to the study website or online Goal Tracker. Figure S1 provides an overview of the trial for both the intervention and control groups.

### Development of the educational information

The nutrition information delivered to both the intervention and control groups was based on the Australian Dietary Guidelines 2013^34^ providing information on body regulation and hunger and replacing nutrient-poor food with nutrient-rich food groups.

Participants in the intervention group also received information about self-compassion that was partially adapted from Neff’s website^38^ and her book^39^, encouraging individuals to treat themselves as a good friend in times of suffering. For the current study, information about self-compassion was modified to address participants’ needs, that is, assisting them with weight-related issues such as body dissatisfaction and a need for self-care. Participants were advised to practice self-compassion every day.

### Goal-setting (intervention group only)

At the beginning of each four-week period, participants in the intervention group were advised to adopt two goals (one for nutrition and one for self-compassion) based on the monthly information they received and track their progress on these goals using the online application over the 12-week study period. A list of goal options was available for each topic to guide participants in setting goals. Participants could choose one of the goal options, modify one of those goals or set their own goals. The investigator also provided monthly feedback on newly set goals by reviewing each participant’s goals and emailing feedback to the participant.

### Statistical analysis

Descriptive statistics were used to describe the baseline characteristics of the study sample. To compare between-group differences in outcome measures at baseline, the Pearson chi-square test was carried out for categorical variables and an independent *t*-test for continuous variables. Fisher’s exact test was used if the Pearson chi-square assumption was violated. Cohen’s *d* effect size was used to describe the magnitude of any differences between two groups, with effect sizes of 0.2, 0.5 and 0.8 representing small, medium and large effects, respectively^40^.

Outcomes were analysed using an ‘intention-to-treat’ approach, meaning that the investigator analysed outcomes for all participants regardless of whether they completed the study ^41^. Missing data were handled using Multiple Imputation (MI). With MI, each missing value is replaced by several random values and consequently, several different completed datasets are generated, these datasets are then pooled into a final result. To generate an imputed dataset sequential regression imputation method was used.^42^

To compare the effect of the intervention between two groups and test whether the changes over time differed between the two groups a series of 2 (time: pre- and post-intervention) x 2 (group: intervention and control) mixed analyses of variance (ANOVAs) were carried out. The interaction term tests whether the change over time is different between the groups. When there was a significant time by group interaction, pairwise comparisons were assessed.

Simple linear regression was conducted to investigate whether the number of days that participants practised self-compassion predicted the level of self-compassion post-intervention. Simple linear regression analyses were also carried out to examine whether the number of times that participants logged into the application or reached their goals predicted total energy intake and total HEQ at post-intervention. All regression analyses were adjusted for the baseline values of the respective outcomes.

Mediation analyses were conducted using the PROCESS macro for SPSS^43^ to test whether the effect of the intervention (group) on the outcome variables was mediated by self-compassion values at post-intervention. Baseline values were included as covariates in the mediation analyses. The mediation analysis generates indirect (mediating) effects and CIs using bootstrapping from the data. Bootstrapping with 10,000 resamples and 95% CIs was used to determine the significance of indirect effects. All statistical tests were two-tailed and differences were considered to be statistically significant at *p*<0.05. Data analysis was performed using Statistical Package for the Social Sciences, version 22 (SPSS Inc, Chicago, III) unless otherwise specified.

## RESULTS

### Response rate and participant characteristics

Of 255 people who completed the initial screening survey 225 met the eligibility criteria and were randomly allocated to the intervention or control group. To conceal the group allocation to participants, interested people received the consent form (relevant to their assigned condition) after randomisation. Since randomisation occurred prior to the consent, the participation rates were different for each group as an uneven number of participants declined to participate in each group (see Figure 2 for the recruitment process). A total of 117 participants consented to participate and were included in the analyses. Of those 74 participants, 35 in the intervention group and 39 in the control group completed the study.

**Figure 2.**
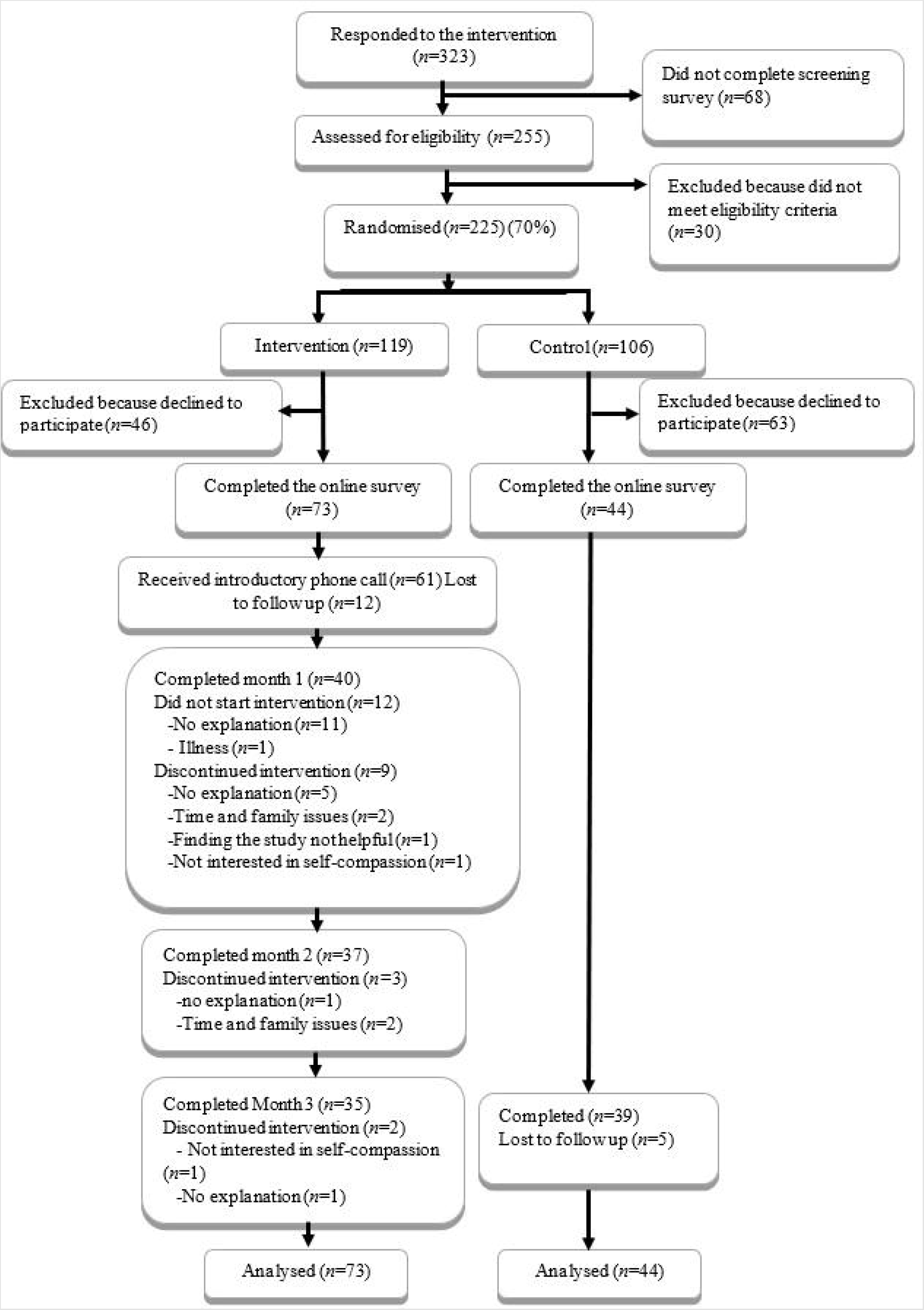
CONSORT diagram of participant recruitment process

Therefore, the retention rate (after consent) was 48% for the intervention group and 89% for the control group. Demographic characteristics were not significantly different between participants who completed the study (*n*=74) and those who did not ([*n*=43], data not shown). Demographic characteristics did not differ significantly between the intervention group and the control group, *p*s ≥0.08 (see Table 1).

**Table 1.** Demographic characteristics of participants who completed the study by group

| Characteristics | | All<br>*n* | Con<br>*n* | Int<br>*n* | $\chi^2$ or Fisher's<br>Exact test(*df*) | *p* |
| --- | --- | --- | --- | --- | --- | --- |
| Sex |  |  |  |  | 0.17(1) | 0.80 |
|  | Male | 18 | 6 | 12 |  |  |
|  | Female | 99 | 38 | 61 |  |  |
|  | Total | 117 | 44 | 73 |  |  |
| Education |  |  |  |  | 1.5(3)<sup>a</sup> | 0.70 |
|  | High school | 8 | 3 | 5 |  |  |
|  | TAFE, college, apprenticeship | 27 | 8 | 19 |  |  |
|  | University undergraduate | 36 | 13 | 23 |  |  |
|  | University postgraduate | 46 | 20 | 26 |  |  |
|  | Total | 117 | 44 | 73 |  |  |
| Ethnicity |  |  |  |  | 8.4(4)<sup>a</sup> | 0.08 |
|  | Oceanian | 41 | 9 | 32 |  |  |
|  | European | 31 | 13 | 18 |  |  |
|  | African and Middle Eastern | 18 | 8 | 10 |  |  |
|  | Asian | 23 | 11 | 12 |  |  |
|  | Americans | 4 | 3 | 1 |  |  |
|  | Total | 117 | 44 | 73 |  |  |
| Income |  |  |  |  | 0.43(4)<sup>a</sup> | 0.37 |
| | < \$25,000 | 11 | 3 | 8 | | |
| | \$25,000 - \$39,999 | 22 | 10 | 12 | | |
| | \$40,000 - \$59,999 | 14 | 8 | 6 | | |
| | \$60,000 - \$79,999 | 14 | 6 | 8 | | |
| | > \$ 80,000 | 54 | 17 | 37 | | |
|  | Total | 115 | 44 | 71 |  |  |

Con, control group; Int, intervention group
<sup>a</sup> Fisher's Exact test

### Self-compassion

Table 2 provides descriptive statistics (means and *SD*s) for the SCS and each of its subscales at pre- and post-intervention, separately for each group as well as the effects of time and group interactions. Assessment of baseline self-compassion scores revealed no significant group differences on any of the subscales, *t*s < 0.48, *p*s > 0.64. There was no significant time-by-group interaction for any of the scores.

**Table 2.**
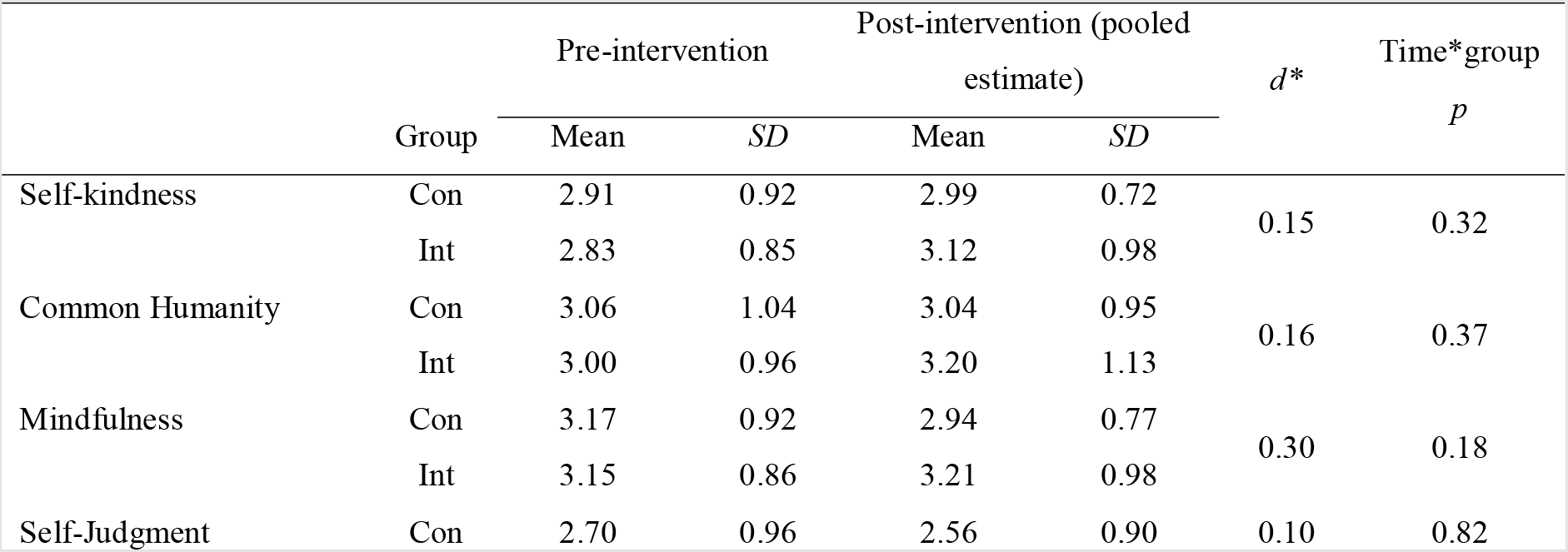
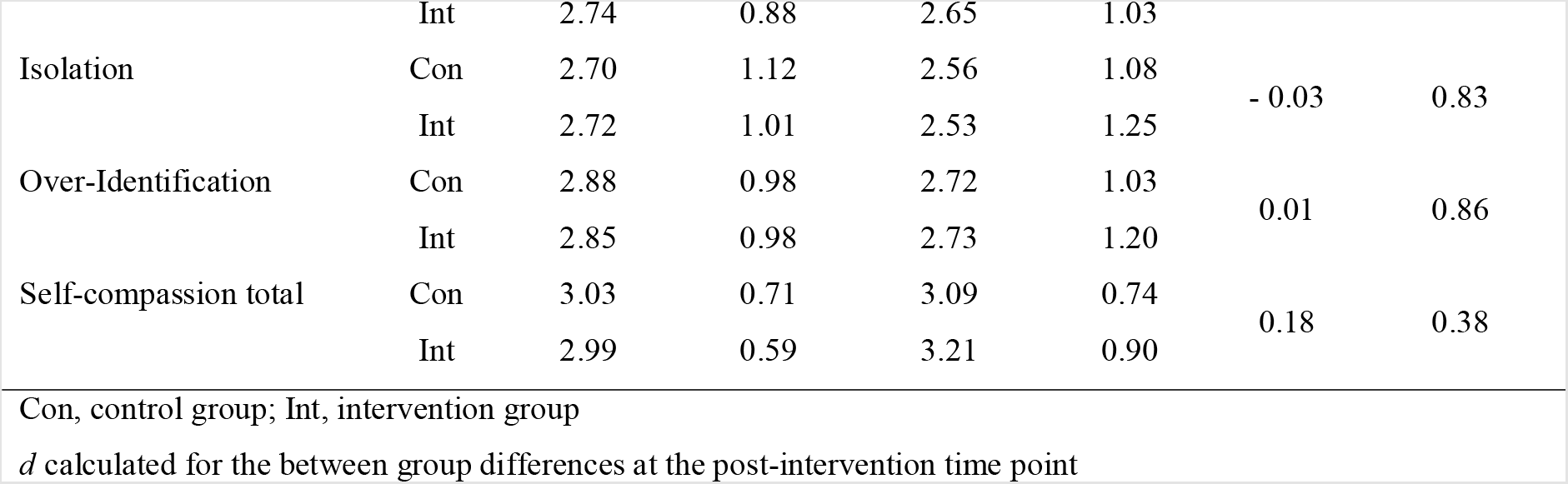
Means, standard deviations and effects of time by group interaction for scores on the Self-compassion Scale for the control (n=44) and intervention (n=73) groups at pre- and post-intervention

### Eating pathology and depression, anxiety and stress

There were no significant differences between the two groups at baseline except for Restraint subscale of EDE-Q: *t*=2.07, *p*=0.04. There was a time-by-group interaction for the Restraint subscale of EDE-Q (Table 3). Pairwise analyses showed that Restraint subscale increased over time in the intervention group (Coefficient=-0.54, *t*=-3.35, *p*=0.002) but did not change in the control group (Coefficient =0.02, *t*=0.09, *p*=0.93). Among the three subscales of DASS, no time-by-group interaction effect was observed.

**Table 3.** Means, standard deviations and effects of time by group interaction for scores on the Eating Disorder Examination Questionnaire and Depression, Anxiety and Stress Scale for the control (n=44) and intervention (n=73) groups at pre- and post-intervention

| | | Pre-intervention | | Post-intervention (pooled estimate) | | $d^*$ | Time*group<br>$p$ |
| --- | --- | --- | --- | --- | --- | --- | --- |
| | Group | Mean | $SD$ | Mean | $SD$ | | |
| EDE-Q |  |  |  |  |  |  |  |
| Restraint | Con | 1.77 | 1.54 | 1.75 | 1.48 | 0.02 | 0.04 |
|  | Int | 1.2 | 1.05 | 1.77 | 1.38 |  |  |
| Eating Concern | Con | 1.32 | 1.15 | 1.02 | 1.06 | 0.18 | 0.29 |
|  | Int | 1.23 | 1.19 | 1.23 | 1.29 |  |  |
| Weight Concern | Con | 3.40 | 1.61 | 2.72 | 1.78 | -0.08 | 0.74 |
|  | Int | 3.33 | 1.31 | 2.60 | 1.36 |  |  |
| Shape Concern | Con | 2.87 | 1.31 | 2.35 | 1.40 | -0.10 | 0.85 |
|  | Int | 2.84 | 1.29 | 2.22 | 1.39 |  |  |
| Global score | Con | 2.34 | 1.11 | 1.97 | 1.20 | -0.03 | 0.50 |
|  | Int | 2.16 | 0.96 | 1.94 | 1.10 |  |  |
| Overeating | Con | 6.15 | 8.21 | 2.93 | 3.93 | 0.10 | 0.90 |
|  | Int | 6.77 | 8.13 | 3.75 | 5.16 |  |  |
| Binging (episodes) | Con | 3.69 | 4.99 | 3.70 | 5.66 | -0.03 | 0.18 |
|  | Int | 5.36 | 7.62 | 3.50 | 5.34 |  |  |
| Binging (days) | Con | 3.75 | 4.82 | 3.29 | 4.03 | 0.05 | 0.45 |
|  | Int | 4.97 | 7.15 | 3.58 | 4.76 |  |  |
| purging (episodes) | Con | 2.42 | 6.02 | 2.79 | 6.36 | -0.29 | 0.29 |
|  | Int | 2.27 | 6.09 | 1.03 | 4.41 |  |  |
| DASS |  |  |  |  |  |  |  |
| Depression | Con | 4.39 | 4.81 | 3.90 | 4.18 | 0.01 | 0.90 |
|  | Int | 4.53 | 3.90 | 3.95 | 3.37 |  |  |
| Anxiety | Con | 3.61 | 4.011 | 3.15 | 3.67 | -0.1 | 0.66 |
|  | Int | 3.55 | 3.64 | 2.76 | 2.47 |  |  |
| Stress | Con | 6.11 | 5.14 | 5.56 | 4.29 |  |  |
|  | Int | 6.92 | 4.52 | 5.56 | 3.23 | <0.001 | 0.35 |

DASS, Depression Anxiety Stress Scale; EDE-Q, Eating Disorder Examination Questionnaire Con, control group; Int, intervention group \*d calculated for the between group differences at the post-intervention time point

### Dietary intake

Regarding data from the 24-hour dietary recall, the only significant difference between the groups at baseline was for fibre intake *t*=-2.22, *p*=0.03. The time-by-group interaction was significant for average daily energy intake, carbohydrates, and marginally significant (*p*=0.054) for saturated fat (Table 4). Pairwise analyses indicate there was a decrease over time in the intervention group for daily energy intake, Coefficient=2139.23, t=3.64, p<0.001 daily carbohydrate Coefficient=56.22, t=2.82, p=0.006 and daily saturated fat intake Coefficient=10.29, t=3.13, p=0.002; in contrast, there was no change observed in the control group for daily energy intake Coefficient=169.29, t=0.22, p=0.82 daily carbohydrate intake Coefficient=-9.43, t=-0.37, p=0.71 and saturated fat Coefficient= 0.04, t= 0.01, p=0.99. There was no time-by-group interaction for the percent of energy contributed by different macronutrients.

**Table 4.** Means, standard deviations and effects of time by group interaction for dietary intake for the control (n=44) and intervention (n=73) groups at pre- and post-intervention

|  | Group | Pre-intervention |  | Post-intervention (pooled estimate) |  | *d* * | Time*group *p* |
| --- | --- | --- | --- | --- | --- | --- | --- |
|  |  | Mean | *SD* | Mean | *SD* |  |  |
| Energy (kJ) | Con | 7490.16 | 3102.13 | 7320.86 | 3821.92 |  |  |
|  | Int | 9236.68 | 5640.37 | 7097.44 | 2411.22 | -0.05 | 0.04 |
| Protein (g) | Con | 79.68 | 38.99 | 79.78 | 43.18 |  |  |
|  | Int | 85.48 | 41.4 | 71.96 | 25.52 | -0.19 | 0.13 |
| Carbohydrate (g) | Con | 216.77 | 96.4 | 226.20 | 121.84 |  |  |
|  | Int | 277.08 | 195.16 | 220.87 | 73.31 | -0.03 | 0.04 |
| Total sugar (g) | Con | 95.79 | 67.03 | 103.98 | 72.65 |  |  |
|  | Int | 126.08 | 131.29 | 100.83 | 43.51 | -0.03 | 0.13 |
| Non-milk extrinsic<sup>b</sup> sugar (g) | Con | 49.30 | 59.45 | 53.25 | 55.53 |  |  |
|  | Int | 77.42 | 120.8 | 51.52 | 41.05 | -0.02 | 0.13 |
| Intrinsic sugar<sup>c</sup> and milk sugar (g) | Con | 45.36 | 32.05 | 50.26 | 32.37 |  |  |
|  | Int | 47.91 | 25.35 | 47.67 | 15.53 | -0.09 | 0.43 |
| Total fat (g) | Con | 69.97 | 50.31 | 61.44 | 40.16 |  |  |
|  | Int | 85.27 | 59.8 | 60.65 | 35.84 | -0.01 | 0.17 |
| Saturated fat (g) | Con | 1.99 | 6.89 | 1.87 | 4.71 |  |  |
|  | Int | 6.49 | 17.93 | 2.81 | 6.70 | 0.07 | 0.06 |
| Fibre (g) | Con | 12.86 | 8.11 | 12.41 | 6.48 |  |  |
|  | Int | 16.39 | 8.48 | 13.70 | 5.14 | 0.16 | 0.23 |
| Fibre (g/MJ) | Con | 1.73 | 0.77 | 1.83 | 1.01 |  |  |
|  | Int | 1.91 | 0.9 | 2.13 | 1.22 | -0.35 | 0.66 |
| HEQ (maximum score) |  |  |  |  |  |  |  |
| Vegetables (21) | Con | 11.50 | 4.42 | 11.64 | 3.92 |  |  |
|  | Int | 11.52 | 4.33 | 12.36 | 2.98 | 0.16 | 0.39 |
| Fruit (12) | Con | 6.34 | 2.29 | 5.95 | 2.39 |  |  |
|  | Int | 5.44 | 2.55 | 5.96 | 2.17 | 0.00 | 0.06 |
| Meat (7) | Con | 3.07 | 1.63 | 3.27 | 1.45 |  |  |
|  | Int | 2.82 | 1.25 | 2.84 | 1.62 | -0.30 | 0.55 |
| Meat alternatives (6) | Con | 2.34 | 1.24 | 2.52 | 1.17 |  |  |
|  | Int | 2.63 | 1.33 | 3.03 | 1.38 | 0.40 | 0.38 |
| Grains (13) | Con | 5.30 | 1.73 | 5.41 | 2.04 |  |  |
|  | Int | 5.62 | 2.14 | 5.58 | 2.43 | 0.09 | 0.74 |
| Dairy (11) | Con | 4.11 | 2.04 | 4.16 | 1.86 |  |  |
|  | Int | 4.18 | 2.02 | 4.65 | 1.79 | 0.24 | 0.31 |
| HEQ total (73) | Con | 33.89 | 6.89 | 34.09 | 7.74 | 0.24 | 0.16 |
|  | Int | 33.59 | 9.128 | 36.00 | 6.57 |  |  |

Regarding data from the HEQ, time by group interaction was close to being statistically significant p=0.06 for the subscale of fruit. Pairwise comparison for the Fruit subscale indicated that, while the score marginally increased in the intervention group, Coefficient=-0.52, *t*=-1.83, *p*=0.07, the change in the control group was not significant, Coefficient=0.39, *t*=1.07, *p*=0.29.

### BMI

There was no significant difference between the two groups’ BMI at baseline *p*=0.51. The average BMI was 29.60 (*SD*=4.30) in the control group and 30.50 (*SD*=4.33) in the intervention group at baseline. The differences in time by group interaction (Coefficient= - 0.20, *t*=-0.35, *p*=0.73) were not significant.

### Participants’ engagement (intervention group only)

Overall, the study website (containing educational information) was visited 1,490 times during the study. Participants on average set 3.5 goals (*SD*=1.0) for nutrition and 2.6 goals (*SD*=0.8) for self-compassion. They also logged into their online Goal Tracker account 36.4 times (*SD*=19.9) over the 12-week intervention (about three times per week). There was no significant association between goal accomplishments (number of times participants clicked ‘Yes’ for nutrition goals) and participants’ dietary intake coefficient beta<0.14, *p*s>0.35.

### Mediation analyses

There were no significant indirect effects for the main outcome variables, including EDE-Q Global score, DASS subscales, daily energy intake, and HEQ total score.

## DISCUSSION

The current online intervention was effective in improving dietary habits in adults with overweight or obesity, a number of changes were observed in dietary habits from pre- to post-intervention in the intervention group but not in the control group.

A novel aspect of this study was the inclusion of a self-compassion component to the intervention. Changes in the levels of self-compassion were not different between the two groups over time. This lack of change compared to previous studies^4,15,44,45^ could be related to the approaches used in the execution of the intervention. In the present study, the self-compassion intervention was online and self-guided, which might be less engaging than interactive in-person sessions^5,46^ and focused on promoting a self-compassionate attitude towards eating, such as dealing with failure in dietary goals/choices, while the SCS was developed to assess self-compassion in general. Therefore, the potential improvement in self-compassion skills that are relevant to eating in the current study might not be generalised to the general sense of self-compassion measured by the SCS. Developing a scale that could reflect this specific domain could capture the change in that specific domain and its association with study outcomes.

We did not observe any significant difference between the two groups in eating pathology or psychological distress over time ^47^. It is worth noting that, dietary habits improvement in the intervention group did not induce maladaptive eating behaviours (as has been observed in some other weight management studies^48^).

Regarding dietary habits, there were several positive effects of the intervention. The intervention group showed decreases in total energy intake, as well as the intake of carbohydrate, but there was no change over time in the control group. Furthermore, a decrease in saturated fat consumption and improvements in the Fruit scores of the HEQ were observed for the intervention group, but not in the control group. Previous studies had similar findings to the current research all showed promising results. These studies showed that interventions with a self-compassion element increased fruit and vegetable consumption^15,49^ and fibre intake^14^ and decreased the intake of total energy, sugar^14^ and high fat food^15,16^.

Similar to previous studies, changes in the secondary outcomes of the BMI were not significant, which is perhaps not surprising given that the focus was on changing dietary habits rather than restricting calories in effect to lose weight. ^47,16^.

The mediational analysis did not support the hypothesis that changes in self-compassion could account for the effect of the group condition (intervention or control) on the main study outcomes. No change in self-compassion found in the present study (possibly due to the online nature of the study) would make it difficult to detect a mediation effect. Further intervention studies are needed with a specific focus on self-compassion status related to eating behaviour to examine the underlying mechanism for the changes observed in the study outcomes.

While the retention rate in this study was not high, engagement with self-monitoring among those who completed the study (intervention group only) was promising, with participants on average logging their entries three times per week which is considered as an acceptable level of engagement^50,51^. Engagement with self-monitoring in the current study is comparable to other studies that used online tools^30,52,53^, and is higher than studies using paper-based self-monitoring^20^. Note that the lack of an association between study outcomes and self-monitoring engagement observed may be due to the fact that adherence was only measured by the frequency of log-ins and goal achievements. In future research, greater precision in the measurement of engagement could uncover an association with study outcomes.

This study has several limitations that should be acknowledged. In the current study, there was an unanticipated high rate of attrition (52%) in the intervention group (the attrition in the control group was 11%). The average rate of attrition in behaviour change interventions is reported to be 18%^54^. One reason for the high attrition rate is that the current study was delivered completely online. Online studies usually have higher attrition rates (ranging between 20-80%), especially when there is no in-person contact compared to the traditional face-to-face method^55^. Some studies indicate that increasing participants’ engagement by providing support (e.g. online social support) and including some level of in-person counselling might improve retention in online interventions^19^.

Despite the intention-to-treat analysis, this high attrition rate may have affected the study results as the missing values are replaced with randomly generated numbers in this analysis. Thus, the impact of the intervention may have been undermined.

## Conclusion

The present study showed promising results that an online intervention that combined self-compassion with goal-setting and self-monitoring could improve dietary habits. Future studies might want to include some strategies such as providing online counselling to increase participants’ engagement and reduce attrition. In addition, studies with a larger scale and longer duration are needed to support these preliminary findings.

## Data Availability

All data produced in the present study are available upon reasonable request to the authors

## Legend for supplementary material

Figure S1. Overview of the trial for both the intervention and control groups

## Notes

### Competing Interest Statement

The authors have declared no competing interest.

### Clinical Trial

ACTRN12620001270909.

### Funding Statement

This study did not receive any funding

### Author Declarations

Ethics committee/IRB of UNSW Human Research Ethics Committee gave ethical approval (approval number: HC16879) for this work.

