## Supplemental Figure 1 for "The efficacy of an online behavioural intervention for improving dietary habits with a focus on self-compassion, goal-setting and self-monitoring: A randomised controlled trial"

**Supplementary file**

**Intervention group**

**Online screening questionnaire**

- Inclusion/exclusion criteria such as body weight, height and medical conditions
- Contact details such as email, phone number

**Control group**

**Online consent**

**Online baseline questionnaire**

**Month 1 information delivered containing:**

- Orientation to the study
- Instructions to the online application and goal-setting
- Nutrition and self-compassion information along with guided goals

**Month 1 follow-up**

- Feedback on goals

**Month 2 information delivered containing:**

- Nutrition and self-compassion information along with guided goals

**Month 2 follow-up**

- Feedback on new goals
- Feedback on the last month’s progress

**Month 3 follow-up**

- Feedback on new goals
- Feedback on the last month’s progress

**Month 3 information delivered containing:**

- Nutrition and self-compassion information along with guided goals

**Online month 3 questionnaire**

Introductory phone call (10 minutes)

**Month 1 nutrition information emailed**

**Month 2 nutrition information emailed**

**Month 3 nutrition information emailed**

Figure S1. Overview of the trial for both the intervention and control groups
